# Autonomous Agents for Auditable Cardiovascular Artificial Intelligence Development

**DOI:** 10.64898/2026.07.10.26357656

**Authors:** Lovedeep S Dhingra, Bruno Batinica, Ryan B Choi, Philip M Croon, Evangelos K Oikonomou, Rohan Khera

## Abstract

Clinical artificial intelligence (AI) models are usually reported as finished artifacts, but each model reflects a limited human search across a much larger space of architectures, inputs, losses, optimizers, and training recipes. We tested whether autonomous code-writing agents could perform a controlled model-development experiment: proposing and evaluating code changes, and seeking performance gains without new data or human-guided edits. We built two such agents: an Iteration Agent that searches sequentially, keeping the best variant at each step, and an Evolution Agent that searches for variations in parallel using multiple large language models and prioritizes high-performing lineages across generations. In two architecturally distinct AI-enhanced electrocardiography (AI-ECG) models for structural heart disease, agent-optimized variants improved rank discrimination across held-out, external, and cross-institution evaluations, with area under the receiver operating characteristic curve gains of +0.006 to +0.039 (paired *p* < 0.05). At a fixed 90% sensitivity, specificity rose by up to 7.1 percentage points and positive predictive value by up to 4.8 percentage points. The selected code changes were substantive, spanning architecture, representation, and training recipe variations. These findings position autonomous agents as an auditable layer for clinical AI model improvement, provided that candidate selection, external validation, and post-update governance are explicit. We release these agents as an open, reusable toolkit.

---

Clinical artificial intelligence (AI) models are usually reported as finished artifacts, with performance summarized after development and validation.^1–3^ In practice, however, every model reflects a finite set of human choices about architecture, inputs, training strategy, optimization, and evaluation.^4–6^ Once a model is published or prepared for deployment, it is rarely clear whether stronger versions could have been developed from the same data, which alternatives were tried and rejected, or how future improvements should be made in a way that is reproducible, auditable, and clinically safe.

This creates an important gap for medical AI. Clinical models cannot be treated as static indefinitely, because patient populations, devices, workflows, and care environments change over time.^7–10^ At the same time, model development and updating cannot rely on ad hoc human experimentation alone. Each proposed change must preserve the clinical task, avoid hidden changes to the data or labels, be tested against prespecified criteria, and leave a clear record of what was changed, why it was changed, and whether it improved or degraded performance.^7–10^ A practical system for clinical AI development therefore needs to do more than search for higher accuracy. It needs to support transparent model improvement within explicit data and validation constraints.

Large language models can now be configured as autonomous coding agents that propose, write, train, evaluate, and revise model code through repeated experimentation.^11–13^ These systems make it possible to search a much larger space of model-development choices than a human team can usually explore manually, including changes to architecture, representation, loss functions, optimizers, regularization, and training recipes.^4–6^ For clinical AI, the key opportunity is not simply automated optimization. If each attempted change is recorded and evaluated under fixed rules, autonomous agents could provide an auditable layer for developing and revising models while keeping the task, data, labels, and validation structure fixed.

AI-enhanced electrocardiography (AI-ECG) provides a useful exemplar for this problem. AI-ECG has become one of the most active areas of cardiovascular AI, with models developed to identify structural heart disease, ventricular dysfunction, hypertrophic cardiomyopathy, aortic stenosis, cardiac amyloidosis, and other conditions from the 12-lead ECG.^14–20^ Hundreds of AI-ECG studies are now published each year, with most focused on developing a new model for a new clinical task.^21–23^ However, as in other areas of medical AI, these models are rarely revisited through controlled, reproducible, and auditable development workflows after publication or validation.^7–10^

We designed two autonomous agents to test this idea (**Fig 1**). The first, the Iteration Agent, follows a single improvement path, keeping a change only when it improves validation performance and then building on it. The second, the Evolution Agent, evaluates multiple candidate models in parallel and prioritizes higher-performing model lineages across generations.^24^ We framed the study as a controlled model-improvement experiment: each agent received the original code, development data interface, and fixed compute budget, with no new data and no human-guided code edits. The goal was to test whether agents could recover stronger model variants from the same development resources while producing a transparent record of the model changes attempted. We evaluated this approach using two independently developed and previously validated AI-ECG models for structural heart disease: the publicly released EchoNext-mini model and a Yale AI-ECG model for a composite structural heart disease outcome.^17,25,26^ Our objectives were: (i) to evaluate the performance of the agent-optimized models across internal, held-out and external validation; (ii) to characterise what each agent changed and how novel the resulting code was; and (iii) to release both agents as an open, reusable toolkit.

**Fig. 1.**
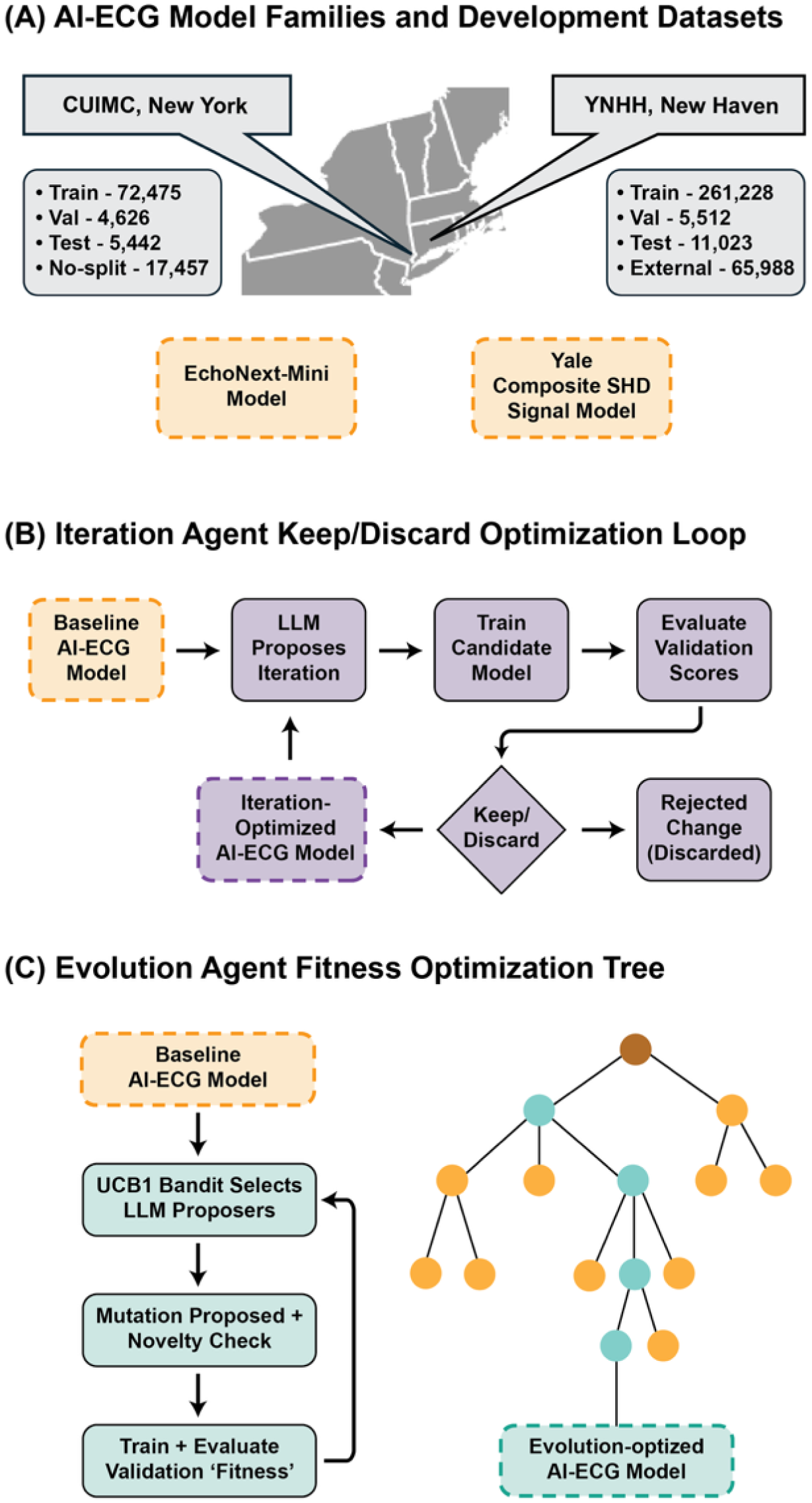
Study design. Abbreviations: AUROC, area under the receiver operating characteristic curve; CNN, convolutional neural network.

## RESULTS

### Performance of agent-optimized models across validation settings

Each agent began with the validated baseline model and ran 100 candidate evaluations. We then selected final optimized models as described in **Methods** and distinguished validation, held-out, external, and cross-institution cohorts according to whether they were used for candidate selection. Both agents improved AUROC for both model families in every evaluated setting, although effect sizes ranged from small to clinically more interpretable operating-point gains. Paired AUROC gains ranged from +0.006 to +0.039 (paired DeLong p < 0.05 for each comparison; **Fig 2**, **Table 1**).

**Fig. 2.**
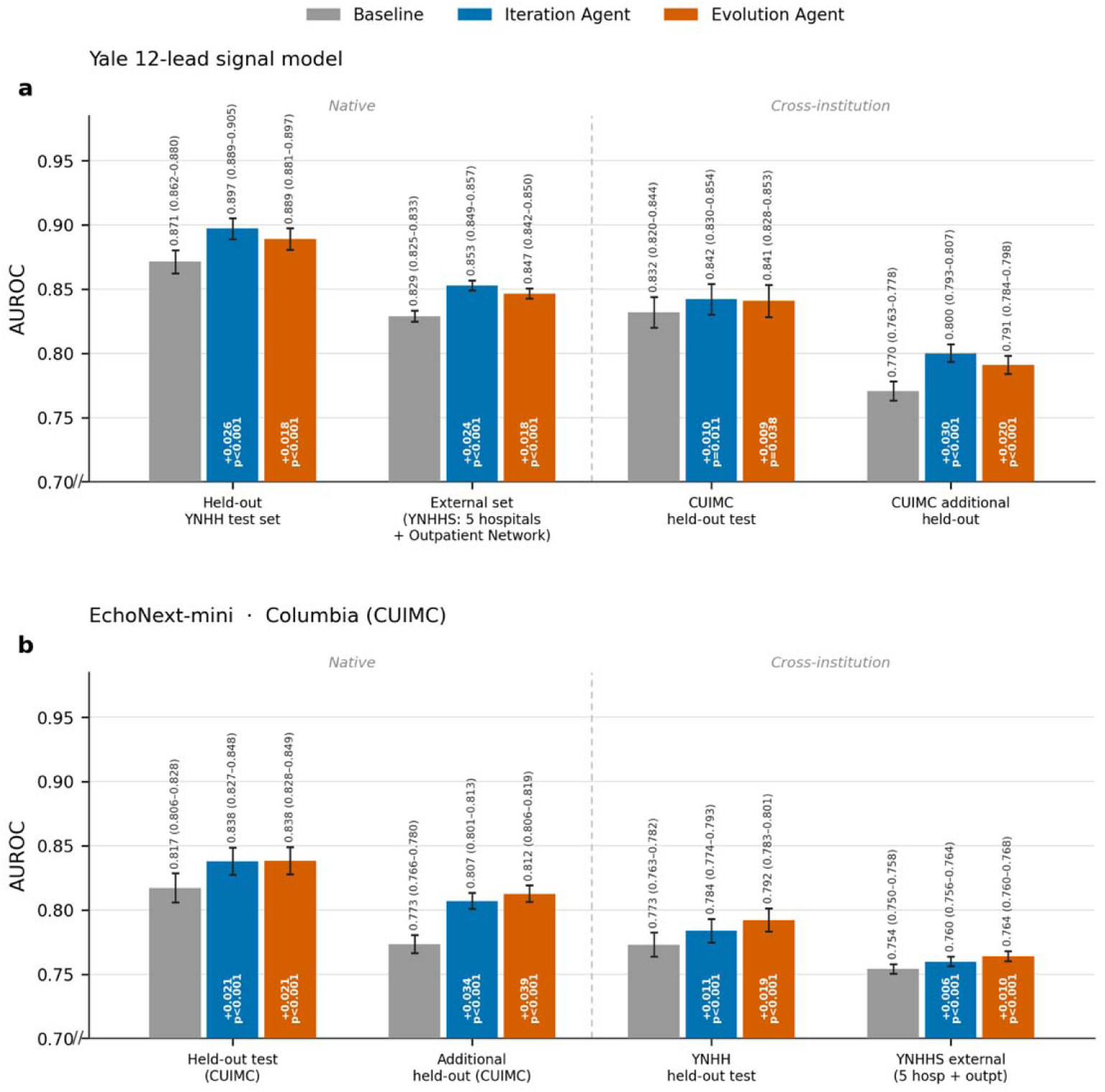
Performance gains across native and cross-institution validation. (a) Yale 12-lead signal model and (b) EchoNext-mini. Each bar is labelled with its AUROC and 95% CI; the gain over baseline and its *p* value are shown within each agent bar. Abbreviations: AUROC, area under the receiver operating characteristic curve; CI, confidence interval.

**Table 1.**
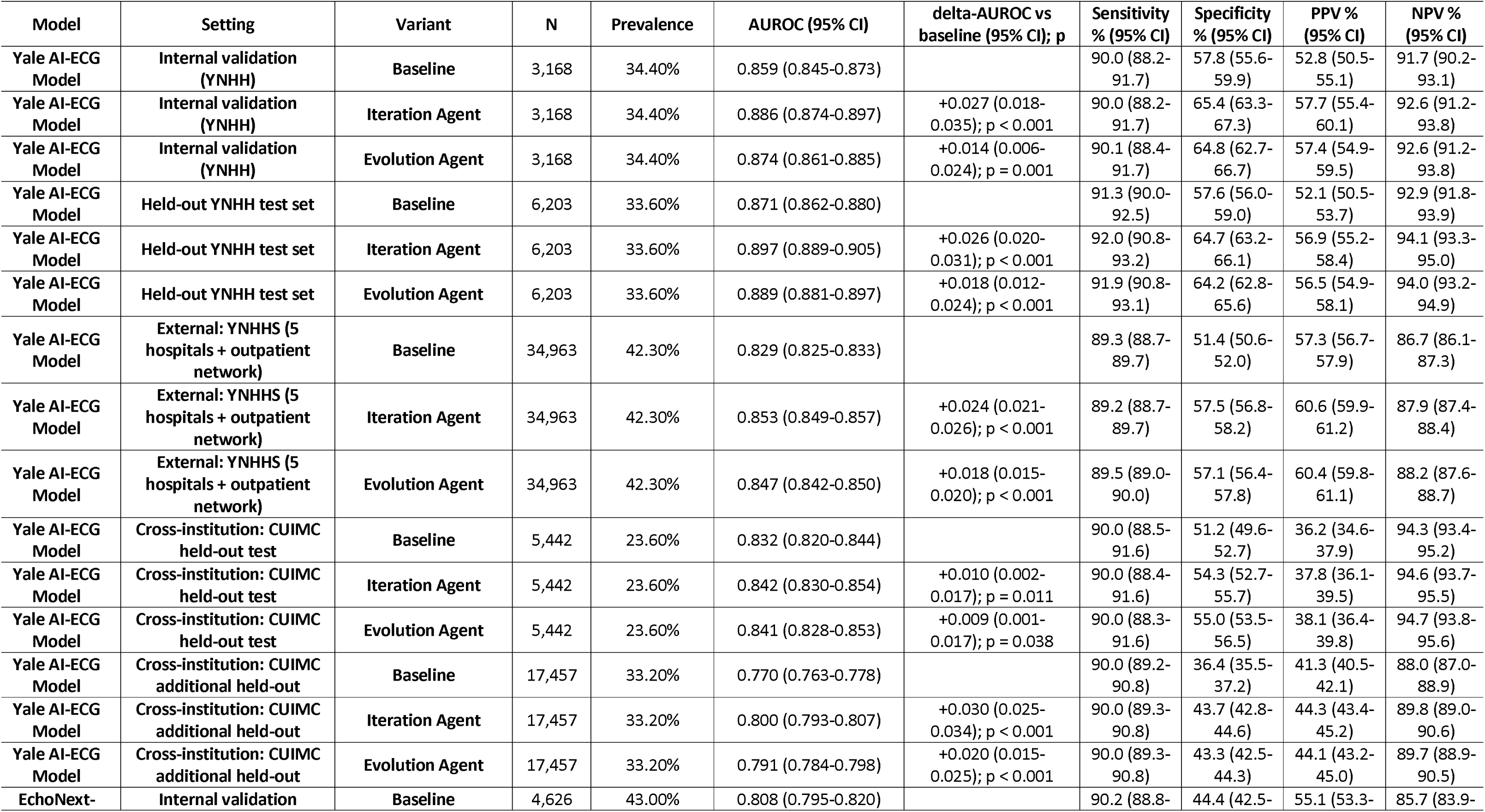
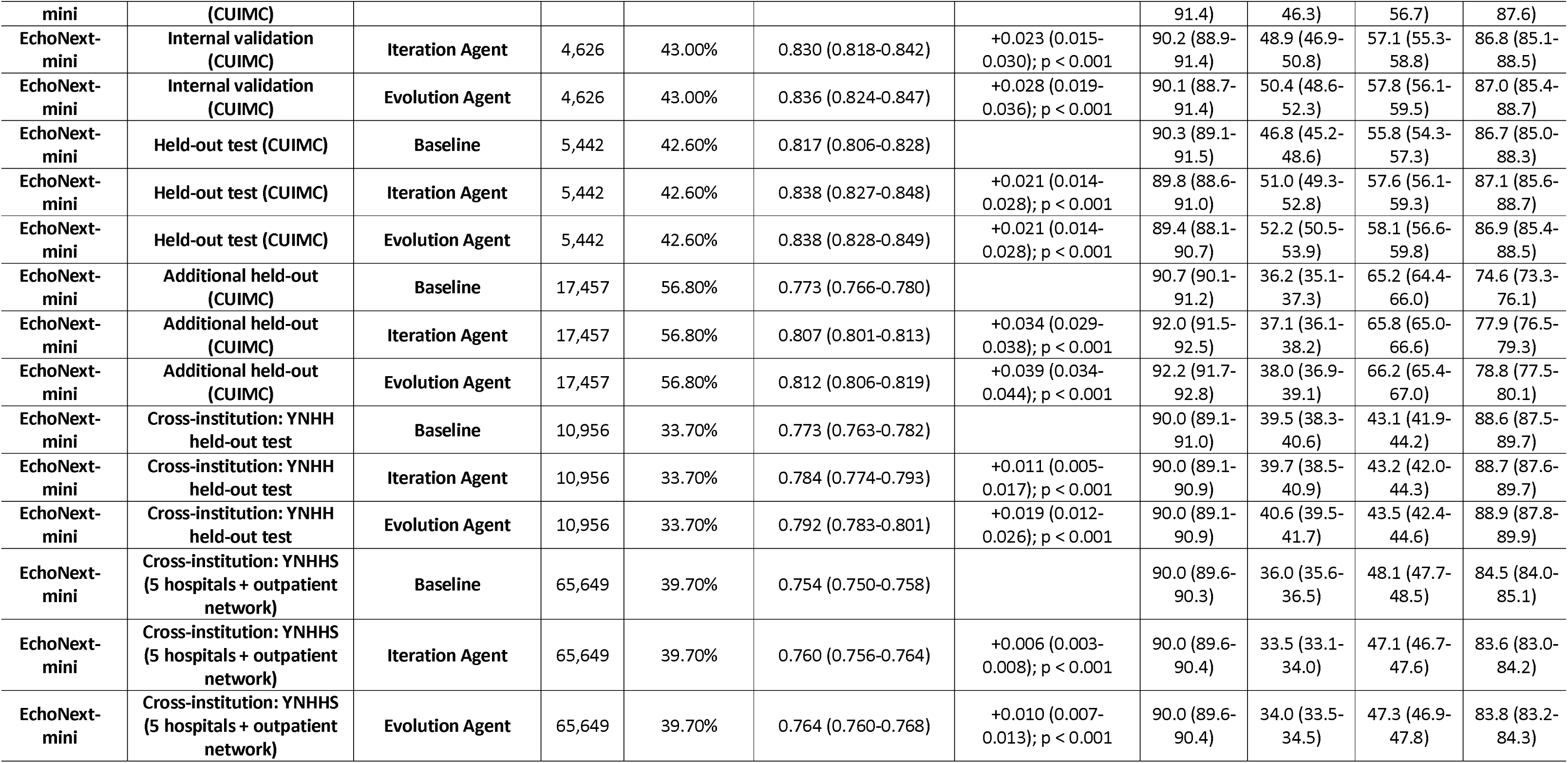
Discrimination and clinical operating points across validation settings.

On the primary held-out test sets, the gains were consistent across the two independently developed model families. The Yale model was evaluated at Yale New Haven Hospital (YNHH; N = 6,203). Its AUROC increased from 0.871 (95% CI, 0.862–0.880) at baseline to 0.897 (95% CI, 0.889–0.905) after Iteration Agent optimization and 0.889 (95% CI, 0.881–0.897) after Evolution Agent optimization. EchoNext-mini was evaluated at Columbia University Irving Medical Center (CUIMC; N = 5,442). Its AUROC increased from 0.817 (95% CI, 0.806–0.828) to 0.838 with either agent. The 95% CIs were 0.827–0.848 for the Iteration Agent and 0.828–0.849 for the Evolution Agent. These gains persisted beyond the primary test sets. On the Yale New Haven Health System external cohort, which included five hospitals and an outpatient network not used for model development, the Yale model improved from an AUROC of 0.829 to 0.853 with the Iteration Agent and 0.847 with the Evolution Agent. On EchoNext-mini’s additional held-out CUIMC cohort, AUROC increased from 0.773 to 0.807 and 0.812, respectively. The optimized models also transferred across institutions, with AUROC gains of +0.006 to +0.030.

We also examined performance at the prespecified screening threshold. For each model, the threshold was chosen at 90% sensitivity in internal validation and applied unchanged elsewhere. Sensitivity remained near 90% across datasets. For the Yale model, specificity increased by 5.8 to 7.1 percentage points and positive predictive value (PPV) by 3.1 to 4.8 points across its native held-out and external cohorts. For EchoNext-mini, specificity increased by 4.2 to 5.4 points and PPV by 1.9 to 2.4 points on its primary test set, with smaller improvements on its additional held-out cohort. Threshold transfer across institutions was less uniform. The Yale model preserved specificity gains of 3.1 to 7.3 points on CUIMC ECGs. EchoNext-mini showed minimal change on the YNHH test set and modest decreases in specificity and PPV on the larger YNHHS cohort. Thus, rank-discrimination gains were more consistent than operating-point gains, emphasizing the need to evaluate thresholds in each intended-use population (**Fig 2**, **Table 1**).

### Characterizing search trajectories for code modifications

To understand how the agents reached these gains, we examined the trajectory of each search and labelled every accepted code change against a predefined, multi-label taxonomy of pipeline components, identifying what kind of change drove each improvement (Methods). The two agentic frameworks followed distinct paths (**Fig 3**): the Iteration Agent advanced along a single improving trajectory, whereas the Evolution Agent branched into several lineages explored in parallel.

**Fig. 3.**
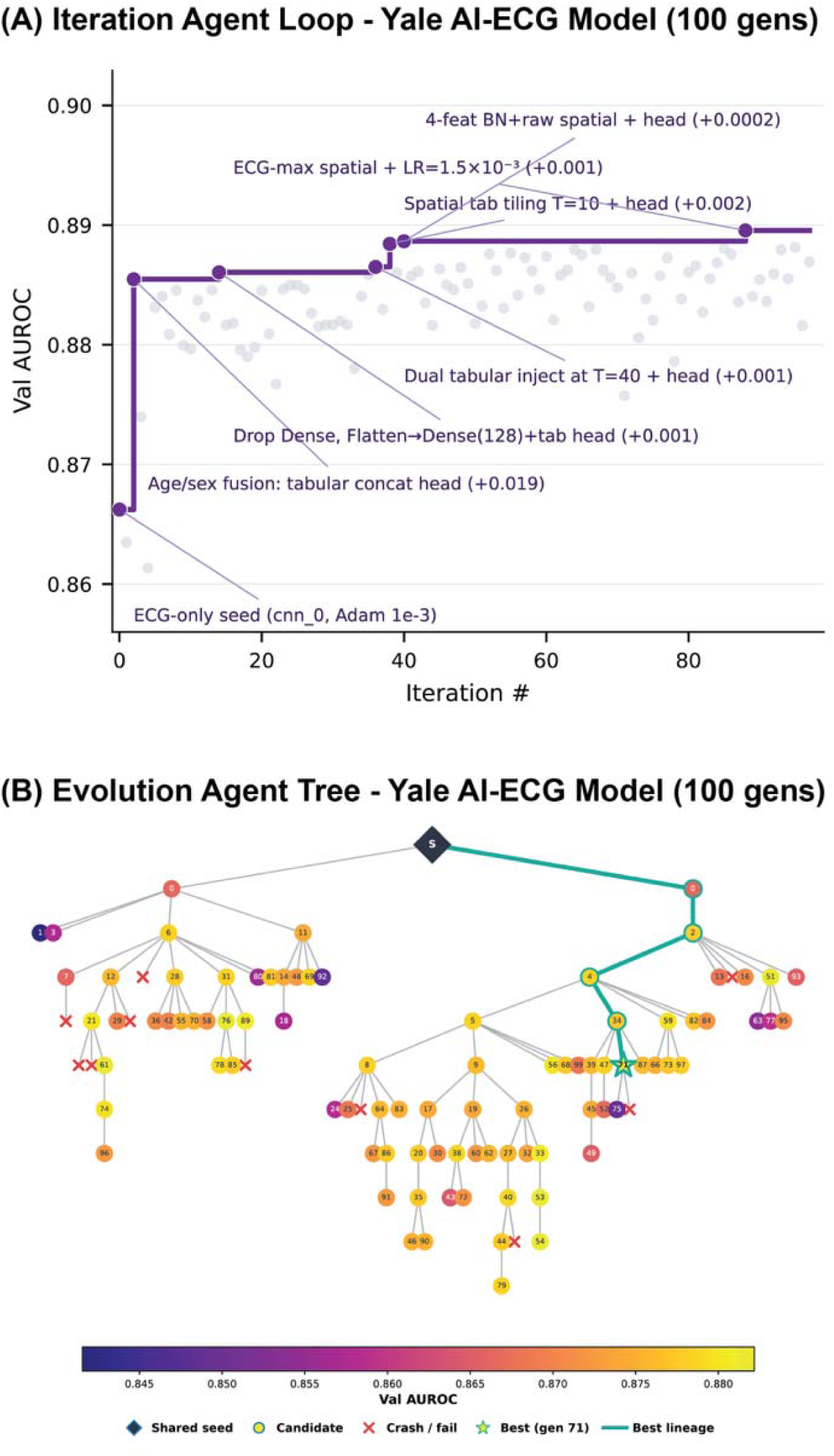
The agentic search process (Yale model). (a) Best-so-far validation score over successive experiments for the Iteration Agent, which follows a single improving trajectory. (b) Lineage tree of the Evolution Agent, in which many variants branch from the shared seed and high-performing lineages are explored preferentially; node colour shows validation AUROC and the highlighted path marks the best lineage. Abbreviations: AUROC, area under the receiver operating characteristic curve.

The composition of accepted changes differed by model family (**Fig 4**). The Evolution Agent changed the model architecture more often than the Iteration Agent in both families: architectural changes appeared in 87% versus 44% of accepted Yale variants and in 30% versus 7% of accepted EchoNext variants. Both agents drew heavily on changes to the loss function, the optimization recipe and regularization, particularly for EchoNext-mini, where these axes were each modified in 24% to 91% of accepted variants and accounted for most accepted changes.

**Fig. 4.**
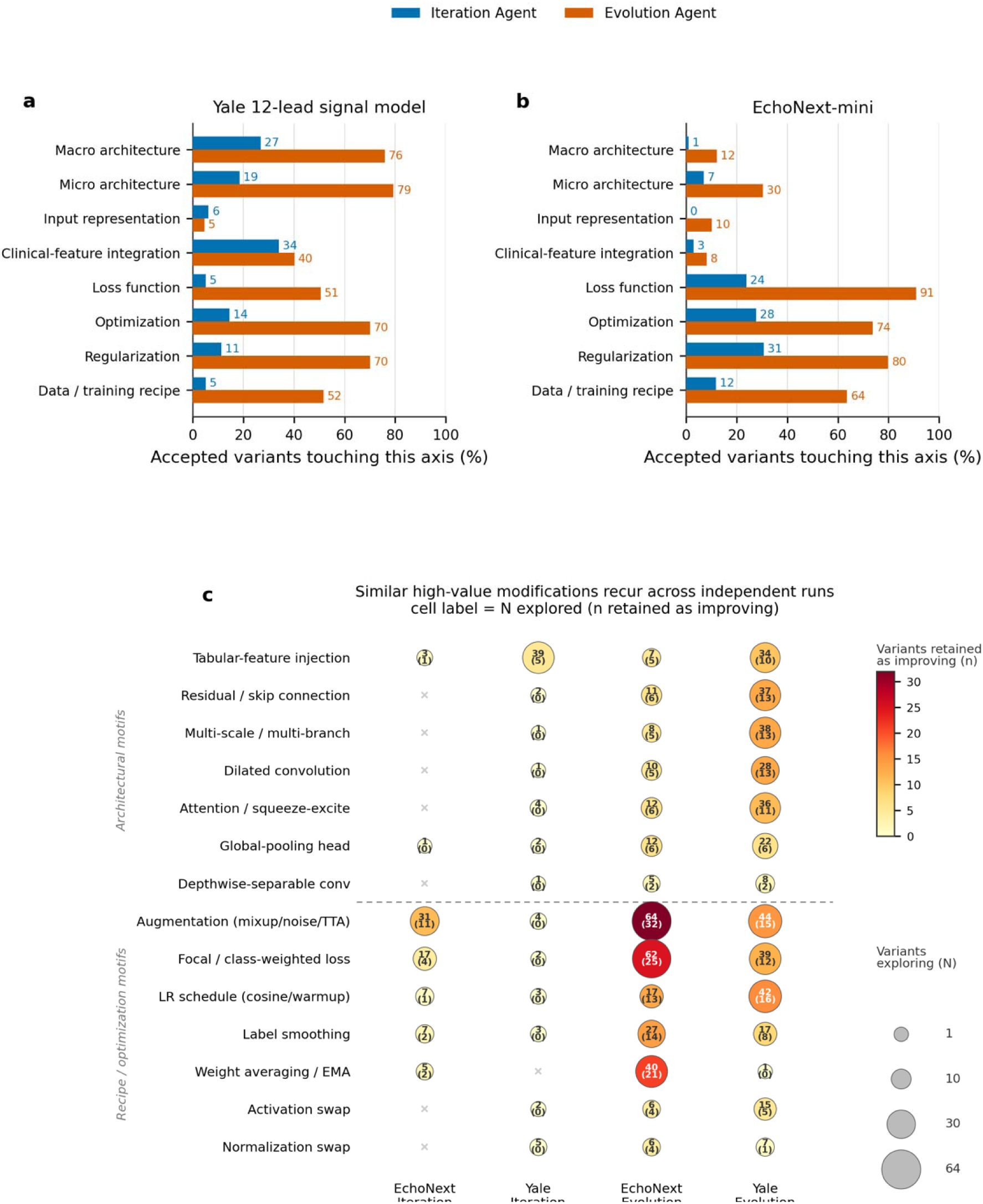
Code modifications by the agents. (a, b) Percentage of accepted variants that modified each pipeline axis, for the Iteration and Evolution agents on the (a) Yale model and (b) EchoNext-mini. (c) Recurring modifications across the four agent-by-model runs; each cell label is the number of variants that explored a motif and, in parentheses, the number retained as improving, with marker size showing the number explored and colour the number retained, separated by an architectural-versus-recipe divider. Abbreviations: EMA, exponential moving average; LR, learning rate; SE, squeeze-and-excite; TTA, test-time augmentation.

Although the agents searched independently, several modifications recurred across all four agent-by-model runs (**Fig 4**). Of the 14 candidate motifs in the taxonomy, six were explored in every run; the four retained most often as improving were data augmentation (58 of 143 variants explored across runs), class-weighted or focal loss (41 of 120), learning-rate scheduling (30 of 69) and label smoothing (24 of 54). Architectural motifs such as residual connections, multi-scale branches, dilated convolutions and attention recurred chiefly within the Evolution Agent but across both model families, showing that the same high-value modifications surfaced repeatedly from independent searches rather than from any single agent or model.

### Embedding-based novelty of code modifications

The taxonomy of accepted changes (**Fig 4**) showed that the agents moved well beyond hyperparameter tuning, altering model architecture, input representation, and the training recipe. To measure the extent of this semantic divergence in code space, we embedded every candidate program in a common text-embedding space and computed each candidate’s divergence from its own agent’s starting point (one minus cosine similarity; **Methods, Fig 5**). The two agents occupied different regions of this space. The Iteration Agent stayed close to its seed, accumulating compact edits, whereas the Evolution Agent explored a markedly wider neighbourhood of code (**Fig 5c, d**). Greater divergence did not come at the cost of performance: in every run the Spearman correlation between a candidate’s divergence and its validation AUROC was non-negative (ρ 0.06 to 0.37). Thus, the most novel variants remained among the strongest.

**Fig. 5.**
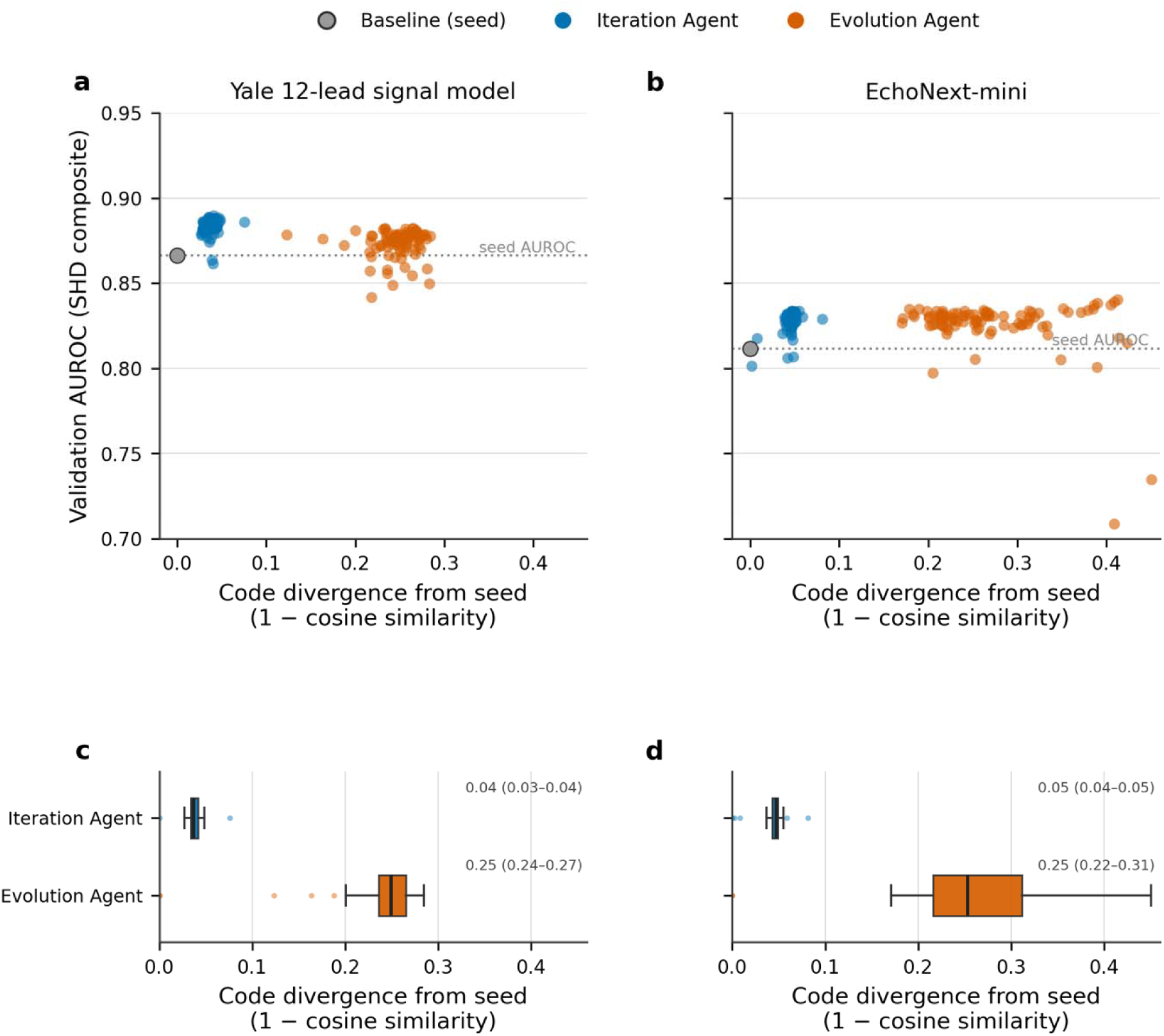
Code novelty. (a, b) Validation AUROC, on a common axis, versus each candidate’s code divergence from its own agent’s starting point (1 minus cosine similarity), for the (a) Yale 12-lead signal model and (b) EchoNext-mini; (c, d) Distribution of divergence from each model’s starting point. Abbreviations: AUROC, area under the receiver operating characteristic curve; IQR, interquartile range.

## DISCUSSION

We applied two autonomous AI research agents to two independently developed and previously validated AI-ECG models for structural heart disease, each working from the original model and development data under a fixed compute budget and without human-guided code edits. Both agents improved rank discrimination in both models across internal, held-out, external, and cross-institution validation; operating-point gains were most consistent in native validation and less uniform in transfer. The agents did more than tune hyperparameters: they rewrote substantial parts of the model code, and several motifs recurred independently across both model families. Thus, validated clinical AI models may contain latent performance headroom that autonomous agents can recover under controlled validation, without additional data.

The central methodological distinction is that the agents edit executable model code rather than only tuning a predeclared hyperparameter space.^4^ Methods such as automated machine learning and neural architecture search tune the choices that developers think to enumerate, within bounds the developers define, and usually constrain the magnitude of each change rather than allowing a model to be restructured.^4–6^ Conventional automated ML and neural architecture search optimize within a human-defined search space. In contrast, autonomous code-writing agents can alter the search space itself by rewriting the training program and reaching design choices the original developers had not explored. Building on this, evolutionary program search driven by large language models has recently been applied to mathematical and machine-learning problems, in frameworks such as AlphaEvolve and ShinkaEvolve.^24,27,28^ Our Evolution Agent is built on ShinkaEvolve, an open-source framework of this kind,^24^ and extends it to a new setting: the autonomous, code-level optimization of clinical AI models that have already been validated, with the resulting gains confirmed on external data.

A notable feature of these results is that headroom remained even in peer-reviewed or internally validated model families. Both improved without any new data, which suggests that at least some carefully built models have performance capacity that systematic code search can still reach. That two independent searches, run on unrelated model families, converged repeatedly on the same kinds of change suggests that these improvements reflect recurring structure in the design space, rather than artefacts of any one model or search.

These agents can be applied at several timepoints in the model’s lifecycle.^7^ A model can be run through an optimization agent before deployment to reach the strongest version its data and available compute can support. After deployment, the agents could be re-run to keep the model current as monitoring detects data and patient populations shift over time. The updated model candidate should be treated as a new model requiring prospective or external revalidation, version control, and governance before clinical use rather than silently replacing the production model. Both uses follow from the same technical capability and the clinical workflow can include change-control and safety review.

The Iteration and Evolution Agents are best understood as complementary search strategies rather than competing ones. The Iteration Agent is a greedy sequential hill-climber that accepts a single improving change at a time, whereas the Evolution Agent is a population-based search that preserves novelty and explores several directions at once.^5,24^ We posit that greedy search may be advantageous when a strong improvement direction exists and can be followed efficiently within a fixed budget, whereas population-based search with diversity preservation may help when the design space rewards broader exploration. In practice, running both paradigms and selecting the better-performing result is a practical default when working under fixed compute.

Because the agents operate by reading code and observing measured performance, the workflow can be portable to any model whose training pipeline can be run and scored, across imaging, signal, and structured-data modalities.^5,24^ To lower the barrier further, we release both agents as code and as plug-in agentic skills for coding environments such as Claude Code and Codex, so groups can apply them to their own pipeline using standard coding-agent infrastructure. The agents are also flexible in what they optimize: their fitness function can be defined by domain experts, and tailored to a clinical priority, favouring sensitivity for a screening tool, specificity for a diagnostic tool, or efficiency in resource-constrained settings.

This study has some limitations that merit consideration. First, each candidate model was evaluated under a fixed time budget rather than trained to convergence. Thus, the reported gains may reflect either better final models or variants that learn faster within the allotted budget. Second, the analysis was retrospective and conducted on existing cohorts. Prospective evaluation will be necessary before the optimized models are used in clinical care. Third, each agent optimized the model’s original composite structural heart disease endpoint, and we did not modify the underlying clinical label definitions or introduce alternative endpoints. We made this choice deliberately so that each agent operated under the same constraints that applied at the time of original development, ensuring that measured gains reflect code changes rather than label redefinition. Fourth, we quantified code novelty using distance in a text-embedding space, which is a useful proxy but does not fully capture programmatic equivalence between variations. However, the taxonomy of changes provides a complementary, human-interpretable view of what the agents altered. Finally, our aim in this study was to characterize the performance gains rather than compute efficiency. A systematic accounting of the cost and efficiency of each agent could help groups judge feasibility and reproduce these searches in practice. Nonetheless, the improvements achieved within fixed time budgets suggest that compute efficiency can be measured empirically rather than treated as a conceptual barrier.

## CONCLUSION

Autonomous research agents improved two validated cardiovascular AI models without new data or human engineering, with gains across held-out and external validation settings. Thus, with transparent selection, compute reporting, and revalidation, agentic workflows could become a reusable strategy for maintaining and improving clinical AI using existing data and compute.

## METHODS

### Models and data

The Yale Institutional Review Board approved the study protocol and waived the need for informed consent, as the study involves secondary analysis of pre-existing, de-identified data. We studied two independently developed and previously validated artificial-intelligence models that detect structural heart disease from the 12-lead electrocardiogram together with a small number of routine structured parameters. The two models were developed on different populations and predict different composite definitions of structural heart disease, and each model was evaluated against the composite label on which it was originally developed. Both models were used in their original deep-learning framework and code, without porting, so that no framework or version differences could confound the comparison.

EchoNext-mini is a publicly released multi-label convolutional model, a one-dimensional residual network with a small branch for tabular inputs, implemented in PyTorch.^17,25^ From a multi-lead waveform and a few routine ECG-derived measurements (including heart rate, PR interval, QRS duration and corrected QT interval), it predicts twelve echocardiographic abnormalities. Its composite structural heart disease label is positive when any of eleven component abnormalities is present: a left-ventricular ejection fraction at or below 45%, a left-ventricular wall thickness at or above 13 mm, moderate or greater aortic stenosis, moderate or greater aortic regurgitation, moderate or greater mitral regurgitation, moderate or greater tricuspid regurgitation, moderate or greater pulmonary regurgitation, moderate or greater right-ventricular systolic dysfunction, a moderate or large pericardial effusion, a pulmonary-artery systolic pressure at or above 45 mmHg, or an elevated tricuspid-regurgitation peak gradient. The model was developed on a single de-identified cohort of 100,000 ECGs from Columbia University Irving Medical Center (CUIMC), released through PhysioNet, partitioned into training (72,475), internal validation (4,626), held-out test (5,442) and an additional held-out set (17,457). The composite label is defined for every record.

The Yale model is a 12-lead signal convolutional network implemented in TensorFlow and Keras that predicts a single composite structural heart disease outcome from the waveform together with age and sex. The composite is positive in the presence of left-ventricular systolic dysfunction (a left-ventricular ejection fraction below 40%), moderate to severe left-sided valvular disease, severe left-ventricular hypertrophy (defined as an interventricular septal thickness greater than 15 mm), or moderate to severe left-ventricular diastolic dysfunction. It was developed at Yale New Haven Hospital (YNHH), with training (261,228) and internal validation (5,512) sets and an internal held-out test set from the same site (11,023), and it was evaluated on a true external set drawn from the Yale New Haven Health System (YNHHS): five hospitals and an outpatient network that did not contribute to development (65,988). Because the composite is defined only where the required echocardiographic components are available, the labelled counts used for evaluation are 3,168 (validation), 6,203 (test) and 34,963 (external).

### Iteration Agent

The Iteration Agent performs a sequential, greedy search over code changes, modelled on an autonomous keep-or-discard research loop. The agent was a large language model (Claude Sonnet 4.6, run at high reasoning effort) operating as a coding agent inside a dedicated, version-controlled workspace. Starting from the original training script as a seed, the agent repeatedly proposed and wrote the modification to the training script, committed the change to version control, trained the resulting candidate under a fixed time budget, and read the validation metric from the run.

Version control serves as the agent’s memory. When a candidate improves on the current best, the agent retains the commit and continues from it, so accumulated changes compound along one improving lineage; when a candidate does not improve, the agent discards it by resetting the workspace to the previous commit, and the failed attempt is recorded in a run log but not carried forward. The agent could alter any part of the training script, including the model architecture, the input representation, the loss function, the optimizer, the learning-rate schedule, data augmentation and the use of structured inputs. It could not change the fixed data interface, the time budget or the random seed, which together define the fitness function and keep candidates comparable. The agent ran without human intervention.

### Evolution Agent

The Evolution Agent performs a parallel, population-based search and is built on an open-source evolutionary program-search framework (ShinkaEvolve).^24^ From the same seed as the Iteration Agent, it maintained a population of code variants organised into islands that evolved semi-independently and periodically exchanged high-performing members through migration. At each generation it selected one or more parents from the population and from an archive of elite programs, and proposed new variants by prompting a pool of large language models (Claude Sonnet 4.6, Claude Haiku 4.5, GPT-5.4 and GPT-5-mini). A proposal took the form of a focused code diff, a full rewrite, or a crossover that combined two parent programs.

Two mechanisms steered the search. A cost-aware bandit (an upper-confidence-bound strategy) learned over the run which language models produced the largest improvements per unit of cost and sampled them accordingly.^29^ A meta-learning step periodically used a separate model (Claude Sonnet 4.6) to summarise which kinds of change had been working, encoding this as written guidance prepended to subsequent proposal prompts Each proposed variant was also checked against a novelty criterion that rejected candidates whose code embeddings were near-duplicates of existing programs in the population. Every proposed variant was trained and evaluated automatically under the same fixed time budget as the Iteration Agent. As with the Iteration Agent, the agent could modify any part of the training script and ran without human guidance.

### Candidate Selection Strategy

Both agents were run from the same seed for 100 candidate evaluations each, under a fixed per-candidate wall-clock training budget so that the search was bounded by compute rather than by hand-tuning. We refer to the seed, evaluated under the same budget, as the baseline (generation 0). The objective that both agents maximised was the validation AUROC of the composite structural heart disease label. For EchoNext-mini the objective additionally subtracted a small penalty for each structured input the model used (0.005 per feature), which rewards models that rely on fewer routine measurements at inference; this is why the best evolved EchoNext model used none of the structured inputs. From each run we selected the top five candidates by validation AUROC, retrained them under the same budget, and evaluated them on the held-out test set. The single best candidate by held-out test AUROC was taken as the final model for that agent and family, and this final model was evaluated on the external set; the final models are identified in Table 1. All evaluations in this study were time-budgeted.

### Change taxonomy and code novelty

To characterise what the agents changed, we defined a predefined, multi-label codebook covering the principal axes of a deep-learning pipeline (model architecture, signal or input representation, feature integration, loss, optimization recipe and regularization), and we additionally flagged whether each change was structural rather than additive. Every accepted code change was compared against its parent to produce a diff, and each diff was labelled against the codebook by a large language model using the batch interface, with a human spot-check of a sample of diffs to confirm the labelling. To assess whether modifications recurred independently, we cross-tabulated a set of recurring motifs across the two agents and the two model families. To quantify how far the agents moved through code space, we embedded the code of every program and variant in a common text-embedding space (text-embedding-3-small) and computed each candidate’s cosine divergence (one minus cosine similarity) from its own agent’s starting program; Iteration Agent variants, recovered from version history, were embedded with the same model so that both agents occupy a single space. We report divergence from each agent’s own starting point rather than from a single shared seed because the two agents begin from different source files (the Evolution Agent edits a compact program whose generation 0 is the published seed, whereas the Iteration Agent edits a full training script), so a common-seed distance would conflate boilerplate differences with genuine exploration. The relationship between divergence and validation AUROC was summarised by a Spearman correlation per model and agent, reported descriptively without significance testing.

### Statistical analysis

Discrimination was summarised by the area under the receiver operating characteristic curve (AUROC), with 95% confidence intervals from 2,000 bootstrap resamples.^30^ For each model we fixed a single decision threshold at 90% sensitivity on the validation set and applied it unchanged to the held-out test and external sets, reporting sensitivity, specificity, positive and negative predictive value, and prevalence at that threshold, each with a bootstrap 95% confidence interval. To compare each evolved model with its baseline, we computed the paired difference in AUROC on the same patients with a paired bootstrap confidence interval and a DeLong test, which avoids inferring significance from the overlap of marginal intervals.^31^

Evaluation beyond the optimization loop comprised two components for each model. The first was each model’s native external or post-development data: the YNHHS external set for the Yale model, and the additional held-out CUIMC set for EchoNext-mini. The second applied each model to the other institution’s ECGs, scored on that model’s own composite structural heart disease outcome. For this cross-institution transfer, each signal was resampled and amplitude-matched to the input convention of the receiving model; preprocessing equivalence was verified by reproducing each model’s published baseline performance on its own data before transfer. Because the two models define structural heart disease differently, the same physical YNHHS external cohort yields different labelled sample sizes under the two composites (34,963 records under the Yale composite, 65,649 under the EchoNext composite, of 65,988 in the full external cohort). For these cross-dataset evaluations, each model’s composite label was constructed on the other dataset from its echocardiographic fields following the same definition used in that model’s original development, and the model was applied to the other dataset’s waveforms after resampling and amplitude-matching each signal to the input convention of the receiving model. We compared each evolved model with its baseline on the new population using the same paired AUROC analysis as for native evaluation.

Statistical tests were two-sided and the significance level was set at p<0.05. Analyses were performed in Python 3.11. Model and agent code are available at [repository URL].

## Data Availability

Individual-level data from the Yale-New Haven Health System cannot be shared publicly due to HIPAA regulations enforced by the Yale Institutional Review Board. The EchoNext data is available on PhysioNet at https://www.physionet.org/content/echonext/1.0.0/.

## ABBREVIATIONS LIST

AI: artificial intelligence
AI-ECG: artificial intelligence-enhanced electrocardiography
AUROC: area under the receiver operating characteristic curve
CI: confidence interval
CNN: convolutional neural network
CUIMC: Columbia University Irving Medical Center
ECG: electrocardiogram
EHR: electronic health record
LLM: large language model
SHD: structural heart disease
YNHHS: Yale-New Haven Health System

## DATA AND CODE AVAILABILITY

Individual-level data from the Yale-New Haven Health System cannot be shared publicly due to HIPAA regulations enforced by the Yale Institutional Review Board. The EchoNext data is available on PhysioNet at https://www.physionet.org/content/echonext/1.0.0/. The image-based AI-ECG models developed at Yale are publicly accessible for research use on the Cardiovascular Data Science (CarDS) Lab website, and the EchoNext-Mini model is available at https://github.com/PierreElias/IntroECG. The study code will be publicly released on GitHub upon publication.

## REFERENCES

1. Khera, R. et al. Transforming cardiovascular care with artificial intelligence: From discovery to practice. J. Am. Coll. Cardiol. 84, 97–114 (2024).

2. Elias, P. et al. Artificial intelligence for cardiovascular care-part 1: Advances: JACC review topic of the week. J. Am. Coll. Cardiol. 83, 2472–2486 (2024).

3. Jain, S. S. et al. Artificial intelligence in cardiovascular care-part 2: Applications: JACC review topic of the week. J. Am. Coll. Cardiol. 83, 2487–2496 (2024).

4. Baratchi, M. et al. Automated machine learning: past, present and future. Artif. Intell. Rev. 57, (2024).

5. White, C. et al. Neural architecture search: Insights from 1000 papers. arXiv [cs.LG] (2023) doi:10.48550/arXiv.2301.08727.

6. Chen, T., Liang, L., Ding, T. & Zharkov, I. Automated Search-Space Generation Neural Architecture Search. arXiv [cs.LG] (2023) doi:10.48550/arXiv.2305.18030.

7. Feng, J. et al. Clinical artificial intelligence quality improvement: towards continual monitoring and updating of AI algorithms in healthcare. NPJ Digit. Med. 5, 66 (2022).

8. Davis, S. E., Embí, P. J. & Matheny, M. E. Sustainable deployment of clinical prediction tools-a 360° approach to model maintenance. J. Am. Med. Inform. Assoc. 31, 1195–1198 (2024).

9. Guo, L. L. et al. Systematic review of approaches to preserve machine learning performance in the presence of temporal dataset shift in clinical medicine. Appl. Clin. Inform. 12, 808–815 (2021).

10. Ötleş, E., et al. Mind the performance gap: Examining dataset shift during prospective validation. arXiv [cs.CY] (2021) doi:10.48550/arXiv.2107.13964.

11. Yang, J. et al. SWE-agent: Agent-Computer Interfaces Enable Automated Software Engineering. arXiv [cs.SE] (2024) doi:10.48550/arXiv.2405.15793.

12. Wang, X. et al. OpenHands: An open platform for AI software developers as generalist agents. arXiv [cs.SE] (2024) doi:10.48550/arXiv.2407.16741.

13. Lu, C., et al. The AI Scientist: Towards fully automated open-ended scientific discovery. *arXiv [cs.AI]* (2024) doi:10.48550/arXiv.2408.06292.

14. Attia, Z. I. et al. Screening for cardiac contractile dysfunction using an artificial intelligence-enabled electrocardiogram. Nat. Med. 25, 70–74 (2019).

15. Ulloa-Cerna, A. E. et al. rECHOmmend: An ECG-Based Machine Learning Approach for Identifying Patients at Increased Risk of Undiagnosed Structural Heart Disease Detectable by Echocardiography. Circulation 146, 36–47 (2022).

16. Cohen-Shelly, M. et al. Electrocardiogram screening for aortic valve stenosis using artificial intelligence. Eur. Heart J. 42, 2885–2896 (2021).

17. Poterucha, T. J. et al. Detecting structural heart disease from electrocardiograms using AI. Nature 1–10 (2025).

18. Veer Sangha, B. S., et al. Ensemble deep learning algorithm for structural heart disease screening using electrocardiographic images: PRESENT SHD. J. Am. Coll. Cardiol. (2025) doi:10.1016/j.jacc.2025.01.030.

19. Dhingra, L. S. et al. Reliability of artificial intelligence-enhanced electrocardiography. medRxiv (2025) doi:10.1101/2025.11.04.25339526.

20. Croon, P. M., Dhingra, L. S., Biswas, D., Oikonomou, E. K. & Khera, R. Phenotypic selectivity of artificial intelligence-enhanced electrocardiography in cardiovascular diagnosis and risk prediction. Circulation (2025) doi:10.1161/CIRCULATIONAHA.125.076279.

21. Wu, Z. & Guo, C. Deep learning and electrocardiography: systematic review of current techniques in cardiovascular disease diagnosis and management. Biomed. Eng. Online 24, 23 (2025).

22. Siontis, K. C., Noseworthy, P. A., Attia, Z. I. & Friedman, P. A. Artificial intelligence-enhanced electrocardiography in cardiovascular disease management. Nat. Rev. Cardiol. 18, 465–478 (2021).

23. Attia, Z. I., Harmon, D. M., Behr, E. R. & Friedman, P. A. Application of artificial intelligence to the electrocardiogram. Eur. Heart J. 42, 4717–4730 (2021).

24. Lange, R. T., Imajuku, Y. & Cetin, E. ShinkaEvolve: Towards open-ended and sample-efficient program evolution. *arXiv [cs.CL]* (2025) doi:10.48550/arXiv.2509.19349.

25. Hughes, J. W. et al. EchoNext-mini: A dataset and baseline AI model for detecting structural heart disease from electrocardiograms. NEJM AI 3, (2026).

26. Dhingra, L. S. et al. Ensemble deep learning algorithm for structural heart disease screening using electrocardiographic images: PRESENT SHD. J. Am. Coll. Cardiol. (2025) doi:10.1016/j.jacc.2025.01.030.

27. Romera-Paredes, B. et al. Mathematical discoveries from program search with large language models. Nature 625, 468–475 (2024).

28. Novikov, A., et al. AlphaEvolve: A coding agent for scientific and algorithmic discovery. arXiv [cs.AI] (2025) doi:10.48550/arXiv.2506.13131.

29. Auer, P., Cesa-Bianchi, N. & Fischer, P. Finite-time analysis of the multiarmed bandit problem. Mach. Learn. 47, 235–256 (2002).

30. Efron, B. & Tibshirani, R. Bootstrap methods for standard errors, confidence intervals, and other measures of statistical accuracy. Stat. Sci. 1, 54–75 (1986).

31. DeLong, E. R., DeLong, D. M. & Clarke-Pearson, D. L. Comparing the areas under two or more correlated receiver operating characteristic curves: a nonparametric approach. Biometrics 44, 837–845 (1988).

